# No clear evidence for relationships of Apolipoprotein E genotype with measures of common infections in three UK cohorts

**DOI:** 10.1101/2024.02.17.24302569

**Authors:** Rebecca E. Green, Alba Fernández-Sanlés, Caterina Felici, Charlotte Warren-Gash, Julia Butt, Tim Waterboer, Marcus Richards, Jonathan M. Schott, Alun D. Hughes, Nish Chaturvedi, Dylan M. Williams

## Abstract

*APOE* genotype is the strongest genetic risk factor for late onset Alzheimer’s disease, with the ε2 and ε4 alleles decreasing and increasing risk relative to the ε3 allele, respectively. Although evidence has been conflicting, several common infections have been associated with Alzheimer’s disease risk, and interactions by *APOE* ε4 carriage have also been reported. Nevertheless, to date, no study has examined relationships between *APOE* genotype and measures of multiple common infections among large population-based studies.

We investigated associations of *APOE* ε2 and ε4 carriage (i.e. non-carrier vs carrier) with serostatus and antibody titers to 14 common pathogens – encompassing herpesviruses, human polyomaviruses, *C.trachomatis, H.pylori*, and *T.gondii* – in three population-based cohorts (UK Biobank, National Survey of Health and Development, Southall and Brent Revisited). Pathogen serostatus was derived using validated antibody cut-offs for relevant antigens and included as an outcome assessing previous infection. Antibody titers were dichotomised among the seropositive subset for each antigen and included as binary outcomes assessing recent immunological responses. We conducted analyses in each cohort using mixed-models, including age, sex and genetic principal components as fixed-effects, and genetic relatedness as a random-effect. In secondary analyses, we additionally assessed i) relationships of *APOE* ε2 and ε4 dosage (i.e. number of copies of the allele of interest), and ii) relationships of *APOE* genotype with continuous antibody titers (rank-based inverse normal transformed). Findings were meta-analysed across cohorts (n=10,059) using random-effects models and corrected for multiple tests using the false discovery rate.

We found no clear evidence of relationships between *APOE* genotype and serostatus or antibody titers to any pathogen, with no strong associations observed in any of our analyses following multiple testing correction. Investigations of *APOE* genotypes with the clinical manifestations of these pathogens, as well as expanding to include other viruses such as SARS-CoV-2, would also be warranted.

## 1. Introduction

Carriage of an *APOE* risk allele is the strongest risk factor for Alzheimer’s disease (AD) other than age (1). The *APOE* gene encodes the glycoprotein apolipoprotein E (apoE), which has essential roles in lipid metabolism and impacts on many biological processes, including immune function (2). There are three main *APOE* alleles – ε2, ε3, and ε4 – defined by genotypes at two genetic variants, and resulting in different amino acids within mature ApoE at residues 112 and 158 (3). These amino acids affect the structure and function of the protein, including receptor binding, affinities for lipoproteins, and stability, as well as disease risk. For example, in comparison to the most common *APOE* genotype (ε3ε3), ε4 carriers have a 3-15 fold increased risk of AD, whereas ε2 carriers have reduced risk (4).

Since several infections have been associated with AD (5), and some evidence additionally suggests that *APOE* genotype may affect susceptibility to or severity of viral infections (3), it is possible the effects of *APOE* on AD risk may be partly mediated through infection status or immunological response. Increased risks of HSV-1, human immunodeficiency virus, and SARS-CoV-2 infection and/or severe outcomes have been reported among ε4 carriers; and risk of seropositivity to hepatitis B and C may be higher among non-ε4 carriers (3,6). While evidence has been conflicting, suggested pathways include different affinities of apoE isoforms for receptors also involved in pathogen entry, such as heparin sulphate proteoglycans and low-density lipoprotein receptors (3,7). However, these studies have been small and investigated few pathogens in parallel, with limited control for confounding. Examining relationships in large cohorts with a wide coverage of pathogen exposure is warranted.

The application of serology panels to large population-based cohort studies such as the UK Biobank (UKB) presents a valuable opportunity to investigate risk factors of infection status and immune response for multiple pathogens in parallel (8). Using these multiplex serology data available in the UKB, as well as two additional population-based UK cohorts - the National Survey of Health and Development (NSHD) and Southall and Brent Revisited (SABRE) – we aimed to investigate associations of *APOE* ε2 and ε4 genotypes with serostatus and antibody titers to 14 common pathogens.

## 2. Materials and Methods

### 2.1 Study design

The study workflow is summarised in Figure 1. As detailed previously (9), we conducted analyses using three population-based UK cohorts: UKB, NSHD, and SABRE. UKB is a large study including >500,000 participants who were approximately were aged 39-73 years at baseline assessments in 2006-10 (10). NSHD is a birth cohort study initially comprised of 5,362 participants born in mainland Britain during one week in March 1946 (11). SABRE is a tri-ethnic study (European, South Asian, and African-Caribbean) including 4,972 participants aged 40-69 at recruitment in 1988-90, stratified by ethnicity, sex, and age (12).

**Figure 1.**
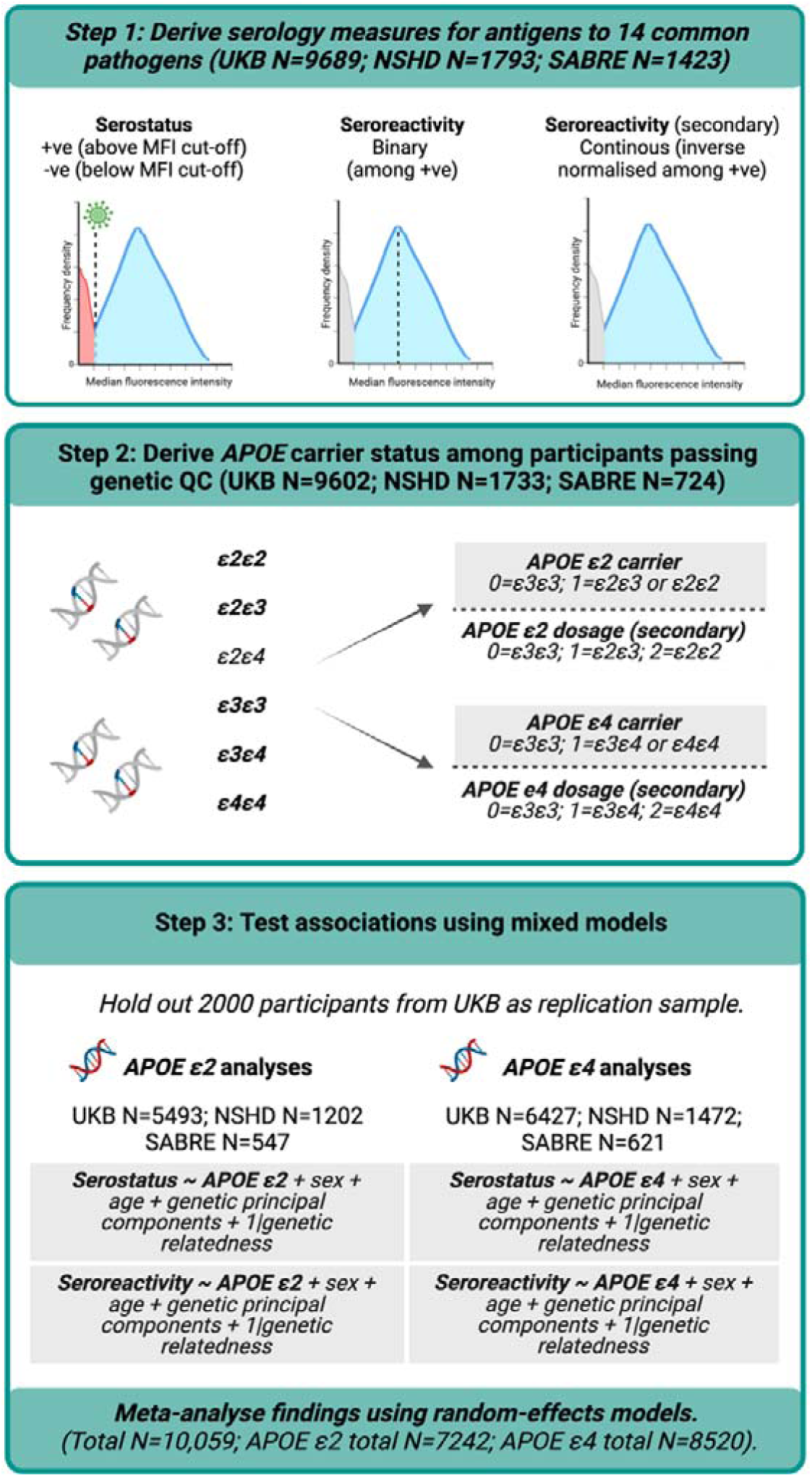
Study workflow. Abbreviations: APOE=Apolipoprotein E genotype, QC=Quality Control. NSHD=National Survey of Health and Development, SABRE=Southall and Brent Revisited, UKB=UK Biobank, +ve=positive, -ve=negative.

All participants provided written informed consent, and all cohorts were granted ethical approval: UKB from NHS North West Research Ethics Committee (11/NW/0382); NSHD from National Research Ethics Service Committee London (14/LO/1173); and SABRE from St Mary’s Hospital Research Ethics Committee (07/H0712/109).

### 2.2 Genetic data and quality control (QC)

Details of genotyping and basic QC of genetic data can be found elsewhere (9,10,13). We used metrics indicative of poor sample quality or sample mix-up to define the analytical subset (excluding those with discordant genetic and self-reported sex, or outliers in heterozygosity and missing rates). We included only biallelic autosomal genetic variants with a call rate >98%.

#### 2.2.1 APOE genotype

*APOE* genotypes were computed using genotypes at two single nucleotide polymorphisms (rs7412 and rs429358), using either directly genotyped or hard-called imputed data. From these, we derived *APOE* ε2 and ε4 carrier status, with carriers defined where participants were heterozygous or homozygous for the allele of interest. We additionally derived *APOE* ε2 and ε4 dosages as a secondary exposure, indicating the number of copies of the allele of interest. In all instances, the non-carrier (for carrier status) or 0 allele (for dosage) groups were comprised only of *APOE* ε3 homozygotes, and individuals with *APOE* ε2ε4 genotypes were omitted (n=272).

#### 2.2.2 Genetic principal components and relatedness

To address a combination of diverse populations and relatedness within samples, we applied PC-AiR and PC-Relate to calculate genetic principal components (PCs) and a kinship matrix (14–16) using the directly genotyped data. PC-AiR calculates genetic PCs using an unrelated and ancestrally representative subset, defined based on cut-offs of genetic relatedness and ancestral divergence (15). These PCs are then projected onto the related subset. PC-Relate estimates relatedness accounting for genetic PCs (16). First, following initial QC detailed in 2.2, genetic data were further filtered for independent common variants (linkage disequilibrium threshold =sqrt(0.1), max sliding window =1×10^-6^, minor allele frequency >0.05). We then performed two rounds of PC-AiR and PC-Relate with the genetic PCs and kinship estimates from the second iteration taken forward for statistical analyses. In round one, kinship was first estimated using KING-robust (17) and genetic PCs were computed by PC-AiR (kinship threshold=2^(−9/2)^ corresponding to 3^rd^ degree relatives; divergence threshold=−2^(−9/2)^). Kinship was then re-estimated using PC-Relate, accounting for these genetic PCs. In round two, genetic PCs were recomputed, this time using the unrelated subset defined using PC-Relate. A kinship matrix was then derived from a second iteration of PC-Relate accounting for these genetic PCs.

### 2.3. Multiplex serology data

Serum immunoglobulin G antibody titers (“seroreactivity”) against a range of antigens for pathogens of interest were quantified using a multiplex serology platform (German Cancer Research Center (DKFZ), Heidelberg), as described previously (9). In brief, 21 pathogens were assayed among 9,689 participants at baseline or instance 1 in the UKB; 18 pathogens among 1,813 NSHD participants at the age 60-64 visit; and the same 18 pathogens among 1,423 SABRE participants at visit 2.

Antigen seroreactivities (expressed in median fluorescence intensity units) were then used to derive pathogen serostatus, based on standardised cut-offs for specific antigens for pathogens. Further details on these measures can be found in Supplementary Notes. We studied pathogens that were measured in all three cohorts with a seroprevalence of >5%. Fourteen pathogens were subsequently included: eight herpesviruses (HSV-1, HSV-2, varicella zoster virus, Epstein Barr virus, cytomegalovirus, human herpesvirus (HHV)-6A, HHV-6B, and HHV-7), three polyomaviruses (JC virus, BK virus, and Merkel Cell virus), two bacteria (*Helicobacter pylori, Chlamydia trachom*)*a*, a*ti*n*s*d the parasite *Toxoplasma gon*. *d ii* As seroreactivities formed a range of non-normal distributions, we derived two measures of antibody response: 1) median split into binary low vs high for that antigen, and 2) rank-based inverse normal transformed variables, which were modelled continuously (secondary outcome).

The two seroreactivity measures were computed among participants who were seropositive to that antigen to avoid including potential noise through antibody cross-reactivity (18). Up to six antigens were quantified per pathogen (Supplementary Notes). Addressing instances where multiple antigens were assayed for a particular pathogen, we used the recommended antigen by DKFZ as our primary outcomes, or randomly selected one (using the R function “sample”) if multiple were recommended.

### 2.4. Statistical analyses

#### 2.4.1 Main analyses

The analytical sample were restricted to those with genetic and serology data. To reduce risk of overinflated estimates due to chance or bias, a replication sample was held out from main analyses (19). We prespecified using a random subset (n=2,000) of UKB participants (selected using the R function “sample”) for replication, rather than NSHD or SABRE; this method was chosen to maximise sample size and similarity with the overall discovery sample.

In our primary analyses, we estimated associations of *APOE* ε2 and ε4 carrier status with i) pathogen serostatus, and ii) antigen seroreactivity (binary) using mixed models. Analytical models were implemented in GENESIS, including genetic PCs, age, sex, and genotyping batch (for UKB only) as fixed-effects, and the PC-Relate kinship matrix as a random-effect. The number of genetic PCs included was chosen based on visual inspection of PC plots. In secondary analyses, we investigated associations of i) *APOE* ε2 and ε4 genotypes with continuous (inverse normal transformed) seroreactivity measures, and ii) *APOE* ε2 and ε4 dosage with the same serostatus and seroreactivity outcomes. The total number of participants differed among analyses because *APOE* ε2 analyses omitted ε4 carriers, ε4 analyses omitted ε2 carriers, and seroreactivity analyses were additionally restricted to the seropositive subset for each antigen.

All analyses were conducted using R. Due to differences in cohort demographics, we ran study-level analyses separately and meta-analysed findings using random-effects models. Significant heterogeneity was defined as I^2^>50% and/or Q-p value<0.05. Each analysis was corrected for multiple tests (within outcome sets, i.e. 14 tests per outcome) using the false discovery rate (FDR; Benjamini-Hochberg method) (20). Findings were defined as statistically significant where p_FDR_<0.05.

#### 2.4.2 Sensitivity analyses

We performed two sensitivity analyses. First, where more than one recommended antigen existed for a pathogen, analyses were repeated using the alternate antigen. Second, we conducted a stratified approach (21), where we restricted analyses to unrelated participants closely clustering with reference panel populations using genetic PCs (see Supplementary Notes). This stratified method can be more robust to possible population structure that may introduce confounding (21,22) but omits many participants, and in our research this only allowed participants closely clustering with “European” reference panel populations to be included. Analyses were conducted using multivariable logistic regression including genetic PCs (derived in our previous work (9)), age, sex, and genotyping batch (for UKB only) as covariates. Findings were meta-analysed using random-effects models.

## 3. Results

### 3.1 Cohort characteristics and seroprevalence

Characteristics of participants included in the present analyses are presented in Table 1. We included participants with genetic and serology data: 9,602 in the UKB, 1,733 in NSHD, and 724 in SABRE (see Figure 1 for participant flow). A subset of the UKB sample (n=2,000) were held out as a replication sample, leaving 7,602 UKB participants for the main analyses. *APOE* ε2 analyses were restricted to non-ε4 carriers (n=7,242), and *APOE* ε4 analyses to non-ε2 carriers (n=8,520).

**Table 1.**
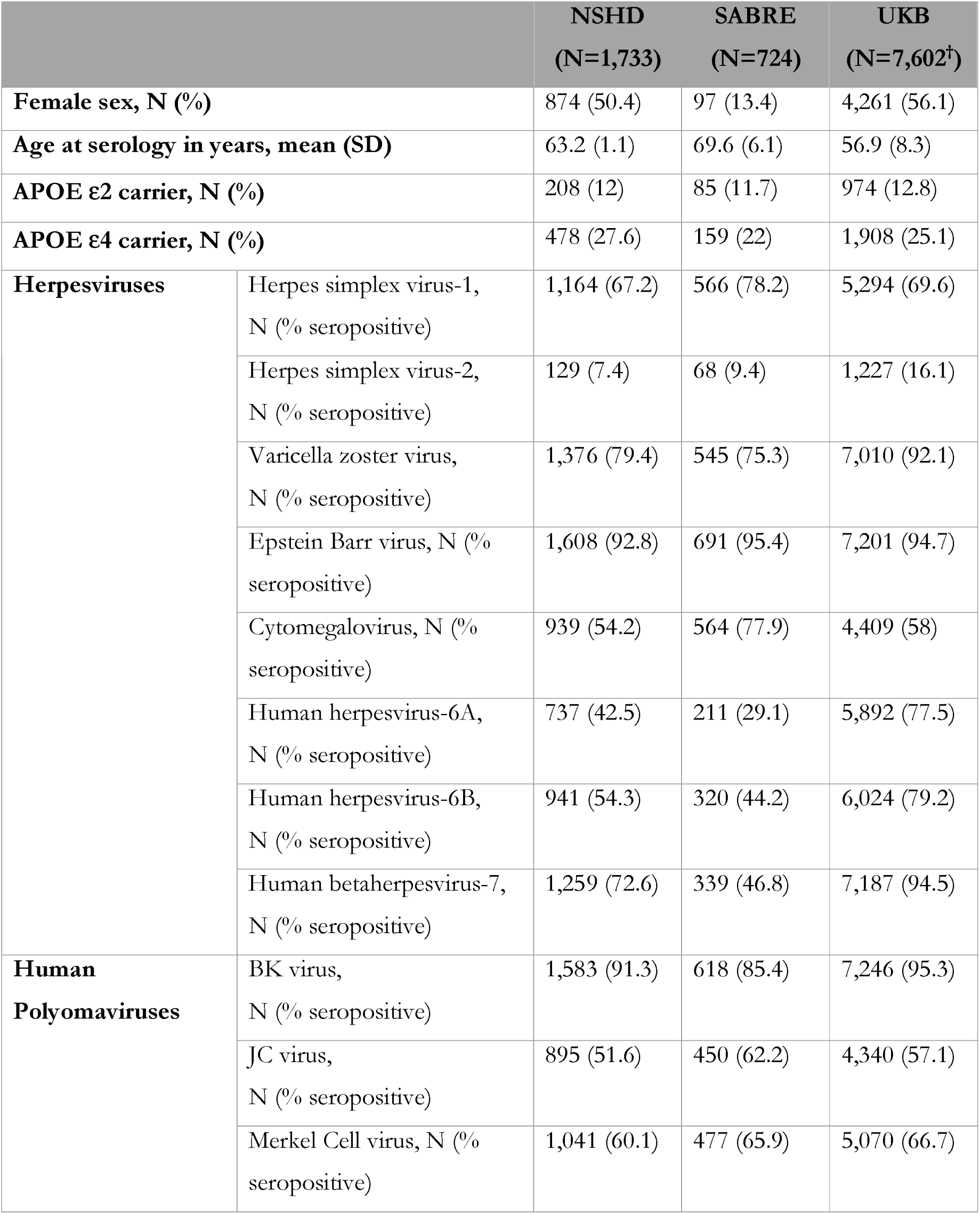
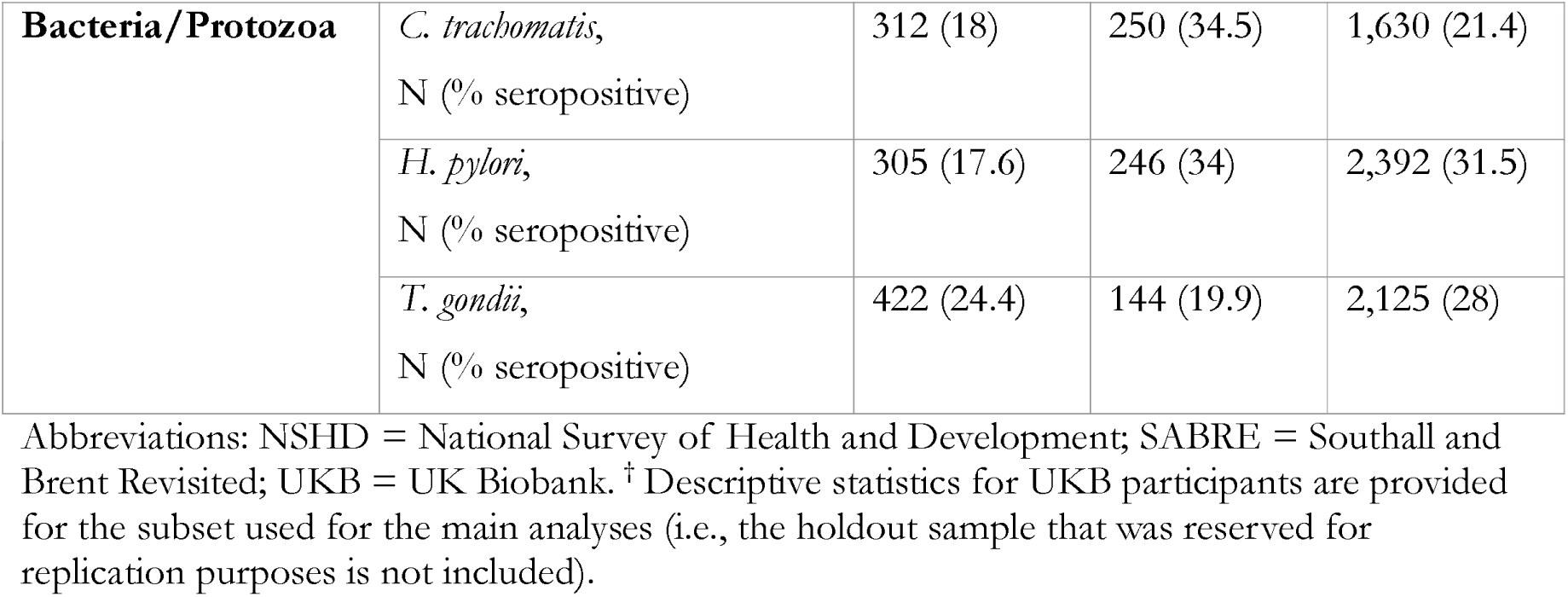
Characteristics of participants with available serology and genetic data.

|  |  | **NSHD**<br>(N=1,733) | **SABRE**<br>(N=724) | **UKB**<br>(N=7,602<sup>†</sup>) |
| --- | --- | --- | --- | --- |
| **Female sex, N (%)** |  | 874 (50.4) | 97 (13.4) | 4,261 (56.1) |
| **Age at serology in years, mean (SD)** |  | 63.2 (1.1) | 69.6 (6.1) | 56.9 (8.3) |
| **APOE $\epsilon 2$ carrier, N (%)** |  | 208 (12) | 85 (11.7) | 974 (12.8) |
| **APOE $\epsilon 4$ carrier, N (%)** |  | 478 (27.6) | 159 (22) | 1,908 (25.1) |
| **Herpesviruses** | Herpes simplex virus-1,<br>N (% seropositive) | 1,164 (67.2) | 566 (78.2) | 5,294 (69.6) |
|  | Herpes simplex virus-2,<br>N (% seropositive) | 129 (7.4) | 68 (9.4) | 1,227 (16.1) |
|  | Varicella zoster virus,<br>N (% seropositive) | 1,376 (79.4) | 545 (75.3) | 7,010 (92.1) |
|  | Epstein Barr virus, N (%<br>seropositive) | 1,608 (92.8) | 691 (95.4) | 7,201 (94.7) |
|  | Cytomegalovirus, N (%<br>seropositive) | 939 (54.2) | 564 (77.9) | 4,409 (58) |
|  | Human herpesvirus-6A,<br>N (% seropositive) | 737 (42.5) | 211 (29.1) | 5,892 (77.5) |
|  | Human herpesvirus-6B,<br>N (% seropositive) | 941 (54.3) | 320 (44.2) | 6,024 (79.2) |
|  | Human betaherpesvirus-7,<br>N (% seropositive) | 1,259 (72.6) | 339 (46.8) | 7,187 (94.5) |
| **Human**<br>**Polyomaviruses** | BK virus,<br>N (% seropositive) | 1,583 (91.3) | 618 (85.4) | 7,246 (95.3) |
|  | JC virus,<br>N (% seropositive) | 895 (51.6) | 450 (62.2) | 4,340 (57.1) |
|  | Merkel Cell virus, N (%<br>seropositive) | 1,041 (60.1) | 477 (65.9) | 5,070 (66.7) |
| **Bacteria/Protozoa** | *C. trachomatis*,<br>N (% seropositive) | 312 (18) | 250 (34.5) | 1,630 (21.4) |
|  | *H. pylori*,<br>N (% seropositive) | 305 (17.6) | 246 (34) | 2,392 (31.5) |
|  | *T. gondii*,<br>N (% seropositive) | 422 (24.4) | 144 (19.9) | 2,125 (28) |

Abbreviations: NSHD = National Survey of Health and Development; SABRE = Southall and Brent Revisited; UKB = UK Biobank. <sup>†</sup> Descriptive statistics for UKB participants are provided for the subset used for the main analyses (i.e., the holdout sample that was reserved for replication purposes is not included).

### 3.2. Main analyses

Results for our main analyses are presented in Figure 2, with full meta-analysed results included in S1-S6 Tables. We report no relationships between *APOE* ε2 or *APOE* ε4 carrier status and serostatus to any of the 14 pathogens in our meta-analyses. While we observed some suggestive associations in our seroreactivity analyses at p<0.05 (namely *APOE* ε2 and antibody levels to *T.gondi*a*i* nd *APOE* ε4 and antibody levels to HHV-6A; see Figure 2 and S1-S3 Tables), no significant associations were reported following multiple testing correction. This was observed both when modelling antigen seroreactivity as a binary variable (low vs. high among the seropositive subset) and in our secondary analyses where values were inverse normal transformed and modelled continuously. We observed similar patterns when modelling *APOE* ε2 and ε4 dosage with these outcomes (S4-S6 Tables). Nevertheless, confidence intervals for some estimates were wide and may not exclude clinically meaningful effects, and we additionally noted instances of significant heterogeneity (I^2^ >50% and/or Q-p value <0.05) in some of these analyses. As no statistically significant relationships were detected after correction for multiple tests, we did not conduct analyses using the pre-specified hold-out replication sample as per our study protocol.

**Figure 2.**
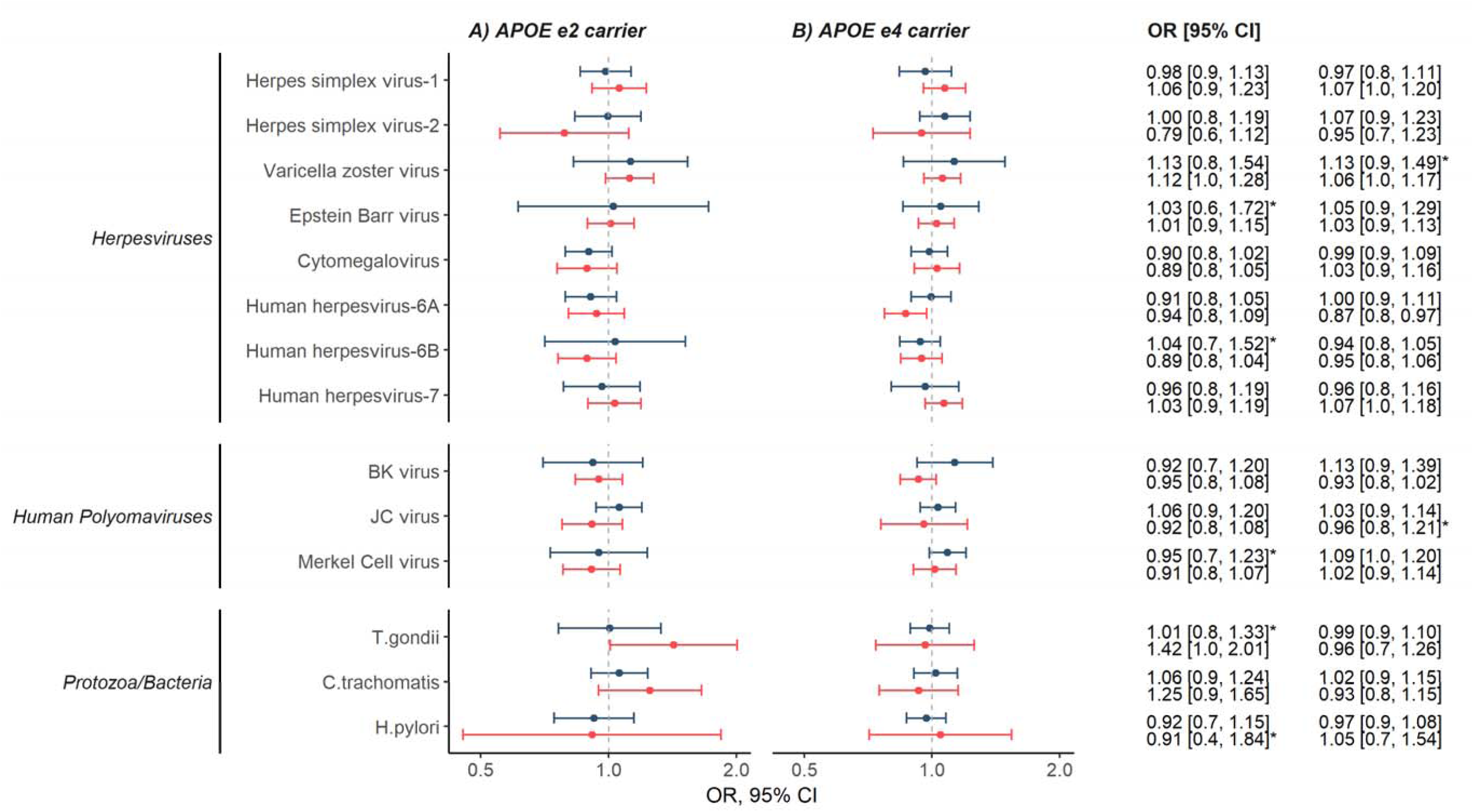
Forest plot and results table indicating our meta-analysed findings of APOE ε2 and ε4 carrier status with serostatus and seroreactivity categories (low vs high). Serostatus results are indicated in blue, and seroreactivity results are plotted in red. Findings where significant heterogeneity (I >50 and/or Q-p value<0.05) were detected are indicated with an asterisk. Seroreactivity analyses were restricted to the seropositive subset only. Total APOE ε2 analysis N=7242, total APOE ε4 analysis N=8520. Full results are available in S1-S6 Tables.

### 3.3. Sensitivity analyses

Results for our sensitivity analyses are included in S7-S9 Tables. No large differences were observed when examining alternate antigens for Epstein Barr virus, cytomegalovirus, and *T.gond*,*i i*nor when conducting analyses restricting to unrelated participants closely clustering with reference panel populations using genetic PCs (N_UKB_ =6,397; N_NSHD_ =1,717; N_SABRE_ =352).

## 4. Discussion

Using three population-based cohorts with genetic and serology data (N=10,059), we investigated associations of *APOE* ε2 and ε4 genotypes with serostatus and seroreactivity to 14 common pathogens (encompassing herpesviruses, human polyomaviruses, *C. trachomatis H. pyl*,*o ri* and *T. gond*)*ii*. This evidence does not suggest common *APOE* genotypes are risk factors for either seropositivity or measures of antibody responses against the pathogens under study, with no clear evidence of relationships observed in all analyses. These null findings were apparent both when modelling ε2 and ε4 carriage and dosage (i.e., number of ε2 or ε4 alleles).

This analysis is the first to assess relationships with many of these pathogens beyond GWAS of the serology measures in UKB (18,23,24). Analyses of the *APOE* locus in GWAS can be complicated through the two single nucleotide polymorphisms encoding *APOE* genotype (rs7412 and rs429358), typically being analysed separately. For example, the minor allele of the variant rs7412 encodes the ε2 allele, and analyses for this variant that assume additive effects (typical for most GWAS) would include ε4ε4 and ε3ε4 carriers in the reference group and a combination of ε2ε3 and ε2ε4 individuals in the heterozygous group. This may be problematic if ε2 and ε4 alleles have different (potentially opposing) effects on a trait, leading to biased results in analyses of both variants. Our study adds to the research base by modelling *APOE* genotype as the combination of these variants, allowing us to examine ε2 and ε4 genotypes separately.

While few studies have evaluated associations of *APOE* genotype with serological measures of infections, associations between *APOE* ε4 and HSV-1 have received most interest, particularly in the context of AD (25,26). Carriage of ε4 has been linked to clinical outcomes such as cold sores following HSV-1 infection (27,28), as well as ε2 with increased risk of herpes simplex encephalitis (29). No strong associations of *APOE* genotypes with antibody titers to HSV-1 were reported in another study, and contrary to our findings here, higher cytomegalovirus antibody titers were observed among ε4 carriers (30). Finally, ε4 homozygosity has been linked to increased risk of shingles (a complication of varicella zoster infection) (7) but this was not supported by another study (31). Nevertheless, these studies were relatively small-scale (max N=1,561) and conducted prior to the increased availability of relevant genetic and infections data. Recommendations for genetic analyses (22), e.g. reducing risk of possible confounding by population structure, were additionally not implemented, increasing risk of false-positive associations.

We did not observe relationships of *APOE* genotype with either serostatus or seroreactivity measures of these pathogens but note that we did not address related clinical outcomes (diagnosed infectious diseases caused by the pathogens under study). In support of our findings here, neither of the two variants conferring *APOE* genotype have emerged as associated loci in GWAS of pathogen serostatus and antigen seroreactivity, as well as infection outcomes such as shingles (18,23,24,32–36). Nevertheless, we note that for some of our estimates confidence intervals were wide, and indeed GWAS may be underpowered for genome-wide scans where a large number of tests are performed. New data releases will present opportunities to broaden our understanding of the genetic architecture of measures of these infections, as well as for other pathogens where *APOE* relationships have been suggested but were not investigated due to low seroprevalence or availability, such as hepatitis B and C (6), and SARS-CoV-2 (37).

Our research has several strengths. It is the largest study to date assessing *APOE* genotype as a risk factor of serological measures of common infections. We used harmonised serology data available in three well-characterised population-based cohorts, where antibody levels to 14 pathogens were available. We additionally included all participants with available genetic and serology data, rather than conducting stratified meta-analyses, where only participants closely clustering with reference panel populations using genetic PCs are analysed, which typically omits many participants and fails to appropriately reflect the continuous nature of genetic variation (21). We also note several limitations. First, while serology measures are able to indicate infection history and immune activity against pathogens without relying on clinical records, they do not inform us of the timing or clinical severity of infection. We were only able to assess associations with antibody titers to pathogens and antigens assayed using the multiplex serology platform, though from our primary motivation of evaluating *APOE* genotypes in relation to pathogens with possible relevance to AD and other forms of dementia, this included several pathogens of interest (principally herpesviruses and other neurotropic pathogens). Second, seroreactivity measures can vary over time (18), and thus individuals may be misclassified into seropositive/seronegative or low/high seroreactivity groups. Third, participants included in these analyses are not fully representative of the wider population; for example, the UKB reports a “healthy volunteer” bias (38,39), and NSHD participants are only broadly representative of those included at recruitment (11). Finally, we may have lacked power for some our analyses, particularly for seroreactivity analyses which are restricted to the seropositive subset for that antigen.

While we did not find evidence for strong relationships of *APOE* genotype with infection status or antibody titers to the 14 infections under study, we present some recommendations for future work. The availability of additional serology data among cohorts with genetic data – as well as the expected expansion of serology data to a larger subset of the UKB – will allow for our analyses to be conducted in larger samples. This may improve the precision of analyses and allow for the inclusion of pathogens that were omitted due to low seroprevalences. Furthermore, interpreting our findings alongside investigations of *APOE* genotypes with clinical outcomes (e.g., through linked primary and/or secondary care records), would provide a more comprehensive view of *APOE* -infection relationships, and additionally permit exploration into infections where serological assays are unavailable.

## Data Availability

Per-study results are available on request.

## Acknowledgements

We are grateful to study participants of the NSHD, SABRE, and UKB. We thank Lee Hamill Howes, Andrew Wong, Kenan Direk, Paulina Januszewicz, and Felicia Huang, as well as other members of the Study Support Team at the MRC Unit for Lifelong Health and Ageing at UCL for their invaluable help towards the generation and curation of NSHD and SABRE data used in this project. This research has been conducted using the UK Biobank Resource under Application Number 71702. JMS acknowledges the support of the National Institute for Health Research University College London Hospitals Biomedical Research Centre, Wolfson Foundation, Alzheimer’s Research UK, Brain Research UK, Weston Brain Institute, Medical Research Council, British Heart Foundation, UK Dementia Research Institute and Alzheimer’s Association. . ADH acknowledges the support of the National Institute for Health Research University College London Hospitals Biomedical Research Centre, Medical Research Council, British Heart Foundation, EU Horizon 2020, Wellcome Trust. We further acknowledge the use of BioRender.com for the creation of Figure 1.

## Abbreviations

AD: Alzheimer’s disease
ApoE: Apolipoprotein E
DKFZ: German Cancer Research Center
FDR: False discovery rate
GWAS: Genome-wide association study
HHV: Human herpesvirus
HSV: Herpes simplex virus
NSHD: National Survey of Health and Development
PC: Principal component
QC: Quality control
SABRE: Southall and Brent Revisited
UKB: UK Biobank

